# Detecting Mental Disorders in Social Media Using a Transformer-Based Ensemble of Binary Classifiers

**DOI:** 10.64898/2025.12.16.25342390

**Authors:** Oleksandr Ovcharuk, Olexander Mazurets, Maryna Molchanova, Alexander Kirpich, Pavel Skums, Olena Sobko, Olexander Barmak, Iurii Krak, Sergiy Yakovlev

## Abstract

This study introduces a novel transformer-based ensemble framework for the multi-label detection of mental health disorders from social media posts. Unlike traditional multi-class approaches that often struggle with comorbidity, the proposed method employs a binary relevance strategy using fine-tuned DistilBERT models to identify co-occurring conditions, including depression, anxiety, and narcissistic personality disorder. To address class imbalance and optimize decision boundaries, the framework integrates a composite loss function (focal, dice, and log loss) and utilizes Youden’s *J* statistic for threshold calibration. Validation on textual datasets demonstrates the efficacy of this approach, with an overall *F*_1_-score of 0.930 and AUC values exceeding 0.89. Comparative analysis suggests that decomposing complex diagnostic tasks into independent binary problems significantly reduces inter-class confusion relative to standard multi-class baselines. Furthermore, a qualitative error analysis highlights specific linguistic challenges, such as contextual polarity shifting, metaphorical ambiguity, and colloquial usage, that impact model specificity. The findings demonstrate the potential of the proposed framework as a robust screening tool for online mental health monitoring, while underscoring the necessity of human oversight to mitigate linguistic misinterpretations.

**Author summary:** Mental health disorders such as depression, anxiety, and narcissistic personality disorder represent a major global health challenge. This work proposes a method that employs transformer-based deep learning models to analyze social media posts for mental health assessment. A significant hurdle in automated diagnosis is that these conditions often occur together (comorbidity), whereas many existing Artificial Intelligence (AI) systems are designed to detect only a single disorder at a time. This study proposes a solution using a “multi-label” deep learning framework. Rather than relying on a single multi-class classifier, the approach utilizes an ensemble of specialized binary models, each trained to detect indicators of a specific disorder.

This design reduces classification confusion between clinically similar conditions, such as depression and anxiety. The method was evaluated on publicly available datasets, had an *F*_1_-score of 0.930 which outperformed the existing approaches. The presented approach demonstrated high effectiveness, achieving better separation between clinically similar disorders compared to traditional methods. Crucially, the detailed investigation beyond the standard statistical metrics was performed which looked into specific models mistakes. It was found that, while the presented AI model is highly sensitive, it can be confused by the specifics of the language such as metaphors (e.g., “feeling like a pressure cooker”), negations (e.g., “I am not worried”), and the colloquial clinical terms. These results highlight that AI is a powerful tool which can be used for early screening and continuous monitoring on social media, while it still requires careful calibration and human oversight to distinguish between genuine symptoms and everyday emotional expression.

The findings demonstrate that analyzing social media texts with advanced machine learning techniques can serve as a powerful complementary tool to clinical diagnostics. While not intended to completely replace professional evaluation, the proposed approach can help identify potential risks, promote earlier detection of mental health disorders, support preventive interventions, and ultimately improve access to care.

## Introduction

Mental health conditions such as depression, anxiety, and narcissistic personality disorder represent a major global health challenge, affecting millions of individuals worldwide. However, many cases remain undiagnosed by conventional assessment methods [1–4]. Over the years, various strategies have been developed to diagnose certain mental disorders and risky behaviors, including depression, eating disorders, gambling, and suicidal ideation [5]. One such strategy involves the monitoring of social media content [6]. This approach enables the identification of behavioral trends by analyzing the lexical, semantic, and stylistic characteristics of individuals’ posts. Social networks function as platforms for communication and as environments where people express their experiences, emotional states, and attitudes toward various events, either explicitly or implicitly. Consequently, analyzing textual content to detect signs of psycho-emotional changes can serve as a valuable tool for the early detection of potential mental health disorders [7].

Conventional methods for diagnosing mental disorders, such as clinical interviews and psychometric assessments, are effective but require direct personal interaction with patients. These approaches are often time- and labor-intensive, have limited throughput, and are typically not proactive [8, 9]. In contrast, the analysis of user-generated content in online environments allows for real-time data collection from large and diverse populations, facilitating earlier and more scalable detection of mental health disorders [10].

The other notable advantage of this approach is its ability to reveal hidden behavioral patterns that might be overlooked by traditional assessments. By utilizing natural language processing algorithms and deep learning techniques, it is possible to analyze vast amounts of text data, pinpointing distinctive linguistic features associated with specific mental disorders. This methodology, therefore, paves the way for the development of automated systems for the early detection of mental health issues, which can enhance clinical methods and improve the overall effectiveness of psychodiagnostics [11].

The study of early diagnostic methods for mental disorders using social media text analysis has a well-established history. Previous research has confirmed the existence of statistically significant lexical and semantic patterns that distinguish social media posts associated with various mental health conditions [12]. It has also demonstrated the feasibility of machine learning approaches, supported by psychological theories, to enhance the understanding of cognitive styles and emotional expressions in identifying conditions such as depression, bipolar disorder, anorexia, self-harm and suicidal tendencies, attention deficit hyperactivity disorder (ADHD), and post-traumatic stress disorder (PTSD) [13–16].

A wide range of machine learning techniques and tools has been employed in the development of diagnostic systems for mental health analysis. These include classical methods such as *n*-gram analysis, Support Vector Machines (SVM), and Random Forests [17], as well as established tools like SentiStrength, the NRC Affect Intensity Lexicon [15], and Linguistic Inquiry and Word Count (LIWC) [18].

In the spectrum of modern computational methods, deep learning architectures play a dominant role, particularly Long Short-Term Memory (LSTM) networks, Convolutional Neural Networks (CNN), Extreme Learning Machines (ELM), and Transformers [16, 18, 19]. A summary of key studies is presented in Table 1.

**Table 1.** Overview of deep learning approaches in mental health classification.

| Ref | Disorders | Dataset Used | Dataset Available | Classification Type/Model DL | Main Results |
| --- | --- | --- | --- | --- | --- |
| [16] | Depression, anxiety, bipolar disorder, ADHD, PTSD | Unstructured user data from Reddit (titles and posts) | No | Multiclass / Transformer | $F_1$ score of 89.65%, outperforming state-of-the-art (89.00%) |
| [18] | Suicide notes | Balanced dataset for suicide notes vs. depressed/blog posts; another with neutral text | No | Binary / RNN | $F_1$ score of 88.26% (experiment 1) and 96.1% (experiment 2) |
| [19] | Depression, anxiety, etc. | Large-scale Reddit dataset with 17.5 million posts from 35,606 users (8 disorders + control group) | No | Binary and multiclass (ensembled) / BERT | User-level binary: $F_1$ scores 0.799–0.931; Post-level binary: $F_1$ scores 0.596–0.736; Multiclass: $F_1$ micro 0.2175, macro 0.195 |
| [20] | Suicide ideation | Reddit dataset with 3549 suicide-indicative posts and 3652 non-suicidal posts | No | Binary / LSTM-CNN | Accuracy of 93.8% and $F_1$ score of 92.8% |

Analysis of the presented data illustrates substantial progress in the application of AI to mental health tasks. Contemporary models often achieve accuracy exceeding 90% when classifying specific, isolated disorders. Furthermore, the integration of textual data with demographic information, clinical assessments [17], and visual modalities [21] demonstrates the potential to further enhance the predictive capability of these systems.

However, performance tends to decline significantly in more complex classification scenarios. For instance, the classification of eight mental disorders [19] (major depressive disorder, generalized anxiety disorder, PTSD, obsessive-compulsive disorder, borderline personality disorder, schizophrenia, eating disorders, and bipolar disorder) based on a large Reddit corpus using a BERT model yielded *F*_1_-scores ranging from 0.57 to 0.73 (mean *F*_1_ = 0.645). These findings highlight the inherent difficulty of simultaneously identifying multiple diagnoses and the presence of significant inter-class confusion. In another study [16], a hybrid architecture combining pre-trained transformer models (RoBERTa, BERT, XLNet, ALBERT) with deep learning methods (CNN, LSTM, BiLSTM) was proposed for the multi-class classification of five mental disorders (ADHD, Anxiety, Bipolar Disorder, Depression, and PTSD) using Reddit posts. The best performance based on *F*_1_-score was by the Transformer model with late fusion, reaching an *F*_1_-score of 0.8965 and outperforming the baseline RoBERTa model (*F*_1_ = 0.8441). Despite the strong performance, this study also reported notable inter-class confusion, particularly among Anxiety, Depression, and PTSD.

Despite these summarized achievements, several critical limitations persist in the field. These include the lack of publicly available datasets suitable for modeling comorbidity and the continued dominance of traditional multi-class classification approaches. When the presented study was conducted, the only open resource enabling multi-label classification of five different disorders (including anxiety, depression, and personality disorders) is the “Text Classification” dataset [22, 23]. Consequently, this dataset was selected as the empirical basis for the present work.

The persisting challenge for studies of mental disorders is the scarcity of data for external validation. In particular, the only available resource, the “Depression Reddit Cleaned Dataset” dataset [24], is limited exclusively to data regarding depression, allowing for only partial verification of the proposed models. The absence of published benchmarks for these new datasets precludes a direct comparison of results.

Consequently, the validation of the proposed method’s efficacy is performed by benchmarking against statistical metrics reported in the review of relevant scientific literature.

The main fundamental issue with existing approaches, which is addressed in this work, is their focus on detecting a single disorder, which contradicts the clinical nature of mental illnesses where comorbidity is often inherent. Traditional multi-class models demonstrate limited effectiveness in identifying co-occurring conditions [25]. Therefore, a transition to a multi-label classification paradigm is a necessary step to adequately reflect the complexity of mental disorders. Particular attention must be paid to the problem of defining the optimal classification threshold to ensure a balance between sensitivity and specificity, which is a critical parameter in medical diagnostics. In this work, instead of empirically selecting thresholds, the presented approach proposes a mathematically grounded method for their optimization. In summary, the proposed approach “shifts away” from the concept of a single diagnosis in favor of detecting a spectrum of conditions. The main contributions of this work are as follows:

- **Multi-label Classification Architecture:** A Binary Relevance strategy based on an ensemble of fine-tuned DistilBERT models is applied. This allows for the independent modeling of each disorder, effectively identifying linguistic markers of comorbidity.
- **Decision Threshold Optimization:** To minimize classification errors between phenotypically similar disorders, Youden’s J statistic is integrated, ensuring an optimal balance of sensitivity and specificity.
- **Sampling Strategy:** A method for creating balanced training subsets for each classifier in the ensemble was developed and applied, mitigating the issue of class imbalance.
- **Loss Function Analysis:** A comparative assessment of the efficacy of Focal Loss, Dice Loss, and Log Loss was conducted, resulting in formulated recommendations for training models on imbalanced medical texts.

## Materials and methods

### Background

DistilBERT is an optimized version of the BERT model [26], created through knowledge distillation, which reduces its size and speeds up its operation while preserving the main properties of the original architecture. In particular, the training process uses BERT as a teacher model, ensuring consistency between the output probabilities [27]. More specifically, DistilBERT undergoes pre-training using masked language modeling (MLM) approach, when some of the tokens in the input text are hidden and the model has to restore them. This approach contributes to the formation of context-dependent representations of words and allows for a more generalized understanding of language structure [28].

From the modeling perspective, one of the key objectives of optimization is to maintain the semantic properties of the hidden states produced by the model [29]. To achieve this a loss function based on cosine proximity is employed, which guarantees a similarity between the vector representations of DistilBERT and its original counterpart. This approach facilitates the development of a compact and efficient model that remains effective across various natural language processing tasks, including text classification. This fine-tuning of DistilBERT is an essential step in adapting the model to classify mental disorders based on the textual content produced by users of social media. For that purpose, the model is trained on a specific selected dataset to better generalize information in the relevant subject area. In the end, the fine-tuning approach for each model involves adding a classification layer to the DistilBERT output layer, allowing the model to determine the probability of one of the studied classes of mental disorders in the text.

To quantify the model performance, loss functions are utilized to evaluate the calibration of DistilBERT models as well as the model performance on training and test datasets. More specifically, the loss function for the DistilBERT model is set to its default when using the Trainer class from the Hugging Face library i.e. the model automatically uses the cross-entropy loss function for binary classification tasks, where the goal is to minimize the difference between actual labels and model predictions.

Tokenization, i.e., the process of converting raw text into a format suitable for the model in the form of tokens, in DistilBERT is performed using the WordPiece tokenizer used in the original BERT model [30]. The tokenization process includes the following steps: i) raw text pre-processing (i.e. normalization, lowercase conversion, removal of extra spaces), ii) breaking the text into sub-tokens according to the WordPiece dictionary, and iii) adding special tokens necessary for the model to function correctly. During the tokenization process each token is assigned a unique identifier (ID) from the corresponding dictionary and the tokenizer also reduces the text to a specified maximum length (128 tokens) to ensure a unified input data representation. The other essential element of the performed tokenization is the attention mask i.e. marking tokens as important (1) and filler tokens (0), which tells the model which tokens to consider during processing and which to ignore.

From the perspective of mental disorders, the model’s task is the accurate classification of the corresponding disorders. At the same time, there exist three types of potential classifications: single-label (i.e. binary and multi-class) and multi-label. Among the first two, binary classification assigns an object to one of the two mutually exclusive classes, while multi-class classification assumes that there are more than two classes, but the classes are still mutually exclusive, i.e., an object can only be assigned to one of the classes at a time [31]. These two types of classification are often referred to as single-label classifications (SLC) because only one class label is used when training the classifier. Multi-label classification (MLC) assigns an object to several classes simultaneously, which are not mutually exclusive. At the same time, MLC can be treated as a special case of SLC. To illustrate that the general classification problem should be considered. More specifically, let *X* be a set of *n* objects to be classified where *X* = {*x*_1_, *x*_2_, …, *x*_*n*_} and *Y* be a set of *m* class labels where *Y* = {*y*_1_, *y*_2_, …, *y*_*m*_}. There is an unknown mapping of the set of objects to the set of classes (classifier): *f* ^*∗*^ : *X → Y*, whose pairs of values are already known on the finite training sample of size *k*: (*X, Y*)^*k*^ = {(*x*_1_, *y*_(1)_), (*x*_2_, *y*_(2)_), …, (*x*_*k*_, *y*_(*k*)_)} where *y*_(*i*)_ is the known label from *Y* which corresponds to the object *x*_*i*_ *∈ X* for *i ∈* 1, 2, …, *k*. The goal is to (re)construct this classifier *f* ^*∗*^ : *X → Y* which assigns *y*_*i*_ ∈ *Y* for each object on *x*_*i*_ *∈ X* for *i ∈* 1, 2, …, *k*. If *m* = 2, then the classification is binary, and if *m >* 2, then it is multi-class. The difference for MLC is that the training set will contain several class label fields and an ensemble of classifiers is required: 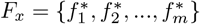, whose dimension is equal to the number of possible class labels (*m*).

There are two main approaches to building MLC classification models: data transformation and algorithm adaptation. The first involves transforming the training dataset, while the second involves adapting conventional machine learning algorithms to create an MLC classifier. The known data transformation methods include binary relevance (BR), classifier chains (CC) and label-powerset (LP). The BR method transforms the MLC problem into several binary classification tasks, each focused on a single label. For each potential class that an object may belong to, a distinct field is created in the training set, where a value of 1 indicates membership in that class, and 0 signifies non-membership. The BR method, however, has two notable drawbacks. The first drawback is the requirement to train multiple classifiers, which can lead to significant computational costs. The second drawback is its failure to consider the interdependence between labels. An alternative approach CC can be used to overcome this label dependence issue of the BR method. The CC technique constructs binary classifiers in a sequential chain, whereby the inputs for each classifier consist of the original objects and the outputs from the preceding classifier in the chain which accounts for interdependence. The LP technique treats every possible combination of labels as a single label in a multi-class problem.

The choice of the metrics for evaluating the accuracy of MLC is crucial. In particular, the selected metrics will vary between conventional classifiers, which predict a single class label, and MLC classifiers, which produce a vector of labels as output. The following specific metrics are often employed to assess the accuracy of MLC classifiers: Hamming loss, which is based on the Hamming distance and defined as the relative frequency of incorrect classifications as well as 0/1-loss, which represents the proportion of training examples for which at least one label has been misclassified. It’s important to note that the Hamming loss metric is typically applied to the BR method while the 0/1-loss metric suits the CC and LP methods. The Hamming Loss metric, however, is not applicable for this study due to the nature of the datasets utilized in the analysis, which are labeled according to SLC tasks. This annotation structure allows for the potential presence of multiple diagnoses for each patient, but only guarantees the presence of a specific disorder associated with the corresponding label. Consequently, using metrics aimed at label completeness, such as Hamming Loss, is unjustified, since the labeling process does not account for the multiplicity of possible labels.

The use of different loss functions such as focal loss, dice loss, and log loss in the presented study is justified by the need to ensure optimal model training in conditions of class imbalance, fuzzy class boundaries, and different sensitivity to classification errors. In particular, the focal loss is a modification of the log loss that inhibits the influence of easily classified examples and focuses training on complex samples [32]. This is crucial in tasks where classes have different expressiveness or are unevenly represented, as in this study. The dice loss function is used in segmentation tasks, but it is effective in classification when the number of examples of the target class is limited [33], since it directly optimizes the *F*_1_- score, which makes it relevant when the balance between precision and recall is important, rather than just prediction accuracy. The log loss (cross-entropy) function remains the standard in binary classification tasks, providing stable convergence and a single benchmark for comparing the performance of alternative loss functions [34]. In summary, the incorporation of all three functions allows evaluating the adaptability of models to different training scenarios and selecting the approaches that best suit the characteristics of the models. In addition to the three discussed losses, it is also useful to evaluate the binary classifiers underlying the BR method using metrics specific to binary classification, which include accuracy – the proportion of correctly classified examples out of the total number; precision – the proportion of correct positive predictions among all predicted positive cases; recall – the proportion of correct positive predictions among all actual positive cases (completeness); as well as the *F*_1_-score – the harmonic mean of precision and recall [35].

The other summaries which were used to evaluate the classifier’s ability to extract hidden dependencies from the training dataset include the visual summaries, such as Receiver Operating Characteristic (ROC) curves and the area under the curve (AUC). In particular, the above indicators are calculated for visual summaries based on the error matrix parameters: True Positive (TP) – the number of correctly predicted positive cases; True Negative (TN) – the number of correctly predicted negative cases; False Positive (FP) – the number of incorrectly predicted positive cases; False Negative (FN) – the number of incorrectly predicted negative instances. Overall, finding an appropriate balance between sensitivity and specificity is particularly relevant for binary classification, where reductions of the FP and FN proportions are necessary. This is especially relevant for medical and psychological research, where correct identification is critical. In this context, the determination of the balance based on the Youden index [36] helps minimize the risk of erroneously assigning text to the mental-disorder category or, conversely, failing to detect it.

### Proposed approach and algorithms

The following notations are used in the method formulations:

- *T* – the input text for analysis. Note that the analysis focuses on user-generated short content from social networks, and therefore, there is a constraint on text length: the number of tokens must not exceed 128.
- *f*_1_ – WordPiece tokenizer [37], which implements the mapping *f*_1_:

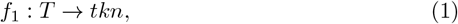

where *tkn* is the tokenised text *T* in the form of a set of tokens;
- *D* – a set of mental disorders identified by *tkn*:

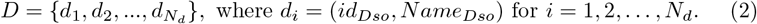

The number *N*_*d*_ denotes the number of mental disorders considered; *id*_*Dso*_ is the identifier of a mental disorder *Dso* (with *id*_*Dso*_ = 0 indicating the absence of any disorder); and *Name*_*Dso*_ represents the name of the corresponding mental disorder.
- *DS* – a dataset with texts are already marked according to their affiliation with a specific mental disorder *d*_*i*_ for *i* = 1, 2, …, *N*_*d*_, which will be used to create datasets for fine-tuning deep learning models:

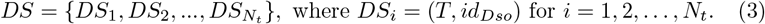

The number *N*_*t*_ is the number of samples in the affiliation *DS*.
- *Df* – a set of datasets for fine-tuning deep learning models for binary classification; each dataset from *Df* contains a set of labeled tokens *tkn* for mapping (1) a specific mental disorder *Cls* = *id*_*Dso*_ or other disorders *Cls* = 0; *Df* is obtained by mapping *f*_2_:

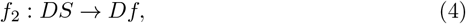

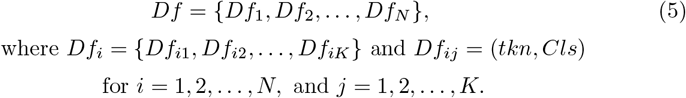

In this notation, *K* denotes the number of samples in the dataset *Df*_*i*_. Each dataset *Df*_*i*_ is constructed for binary classification, where the target class consists of samples related to a specific disorder *d*_*i*_ *∈ D*, and the non-target class includes samples associated with other disorders or with no disorder. To avoid confusion between mental disorders, and considering that a single text may contain manifestations of multiple disorders from the set *D*, the non-target category is formed according to specific rules [38]: (a) the number of samples in the non-target group is equal to, or approximately equal to, the number of samples in the target group, (b) the non-target group consists of an equal proportion of the remaining samples from *DS* that either represent other mental disorders or show no signs of mental disorders.
- *M* – a set of deep learning models obtained using the mapping *f*_3_, which implements fine-tuning of *DLM* using *Df* :

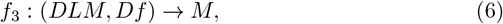

where *DLM* is the DistilBert deep learning model [39] and *M* is a set of finely tuned deep learning models for each mental disorder *d*_*i*_ *∈ D*:

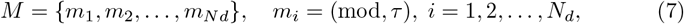

where *mod* is a finely tuned deep learning model *DLM* using the dataset *Df*_*i*_ *∈ Df* for a specific mental disorder *d*_*i*_ *∈ D*;
- *τ* – is a threshold for determining mental disorder *d*_*i*_, which is determined by the Youden index – the value of the point on the ROC curve that corresponds to the best ratio of sensitivity and specificity [40];
- *R* – the result of applying *m*_*i*_ *∈ M, i* = 1, 2, …, *N*_*d*_ to an arbitrary tokenized text *tkn*:

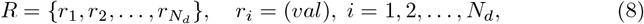

where *val* is the numerical value of the influence of disorder *d*_*i*_ on tokenized text *tkn* with the range of values *r*_*i*_ *∈* [0,1] which indicates the severity of mental disorder *d*_*i*_ *∈ D* [41]. The value of *val* is the output value of the last layer of the model *mod* _*i*_ which is normalized by the *softmax* activation function;
More precisely, *R* is obtained through the mapping *f*_4_:

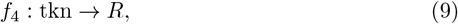

where *R* contains numerical values representing the severity of each disorder *d*_*i*_ *∈ D*. Note that the presence or absence of a mental disorder is determined by comparing the resulting numerical value *val* with the threshold value *τ*.

To provide a complete workflow, Figure 1 presents a diagram of the data processing sequence used to obtain the sets *Df* and *M*. In Figure 1, the first step employs Algorithm 1 to generate a set of datasets *Df* (via the mapping *f*_2_), which are used for fine-tuning a set of deep learning models *M*.

**Fig 1.**
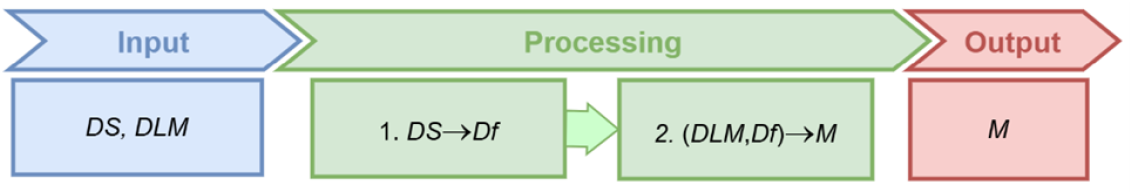
Data processing scheme for obtaining the set of datasets *Df*, which are used to generate deep learning models *M*.

#### Algorithm 1

Implementation of the *f*_2_

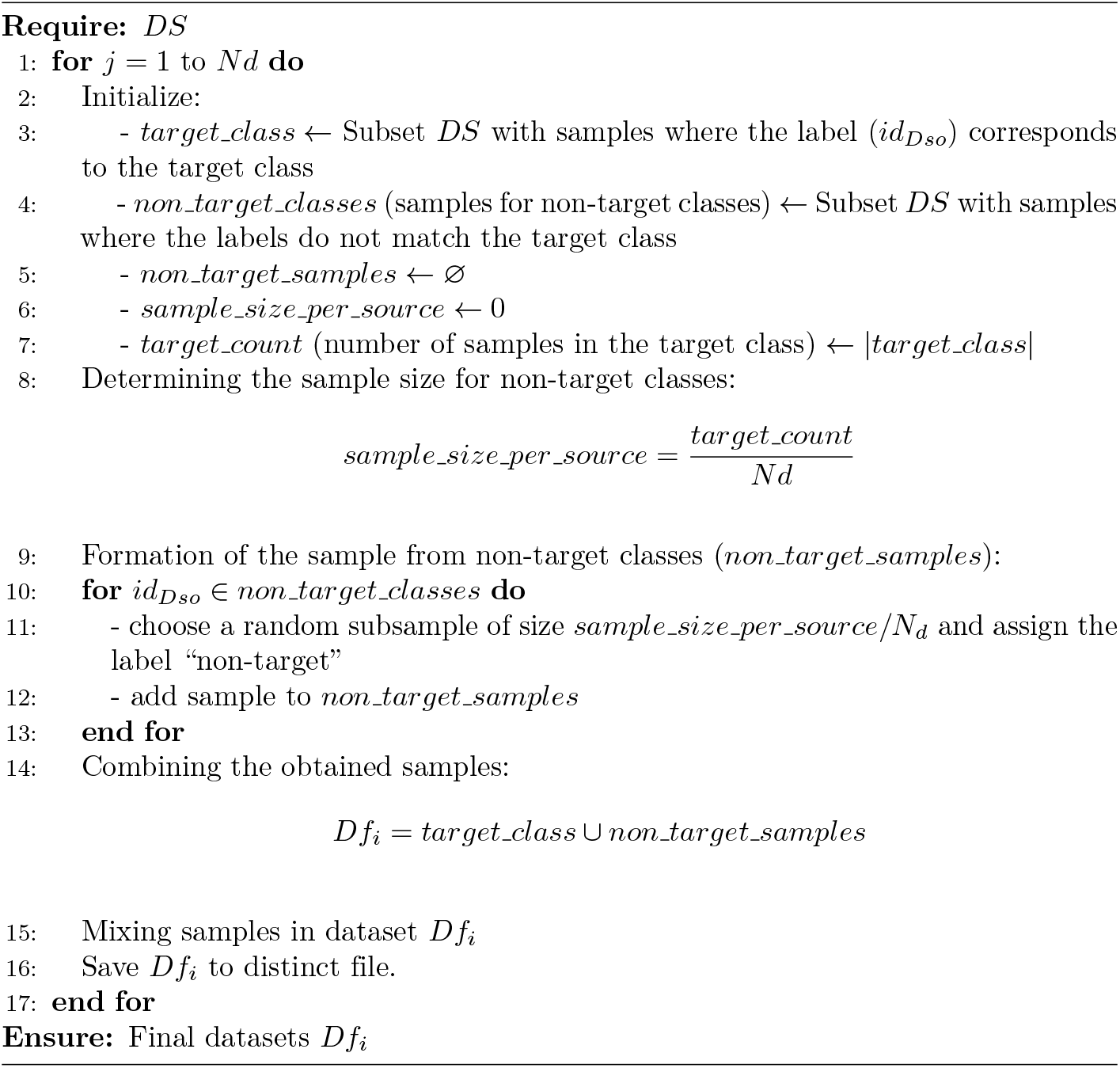

At Step 2 (see Fig. 1), Algorithm 2 is used to fine-tune the set of deep learning models *M* via the mapping *f*_3_.

#### Algorithm 2

Fine-tuning models *M*

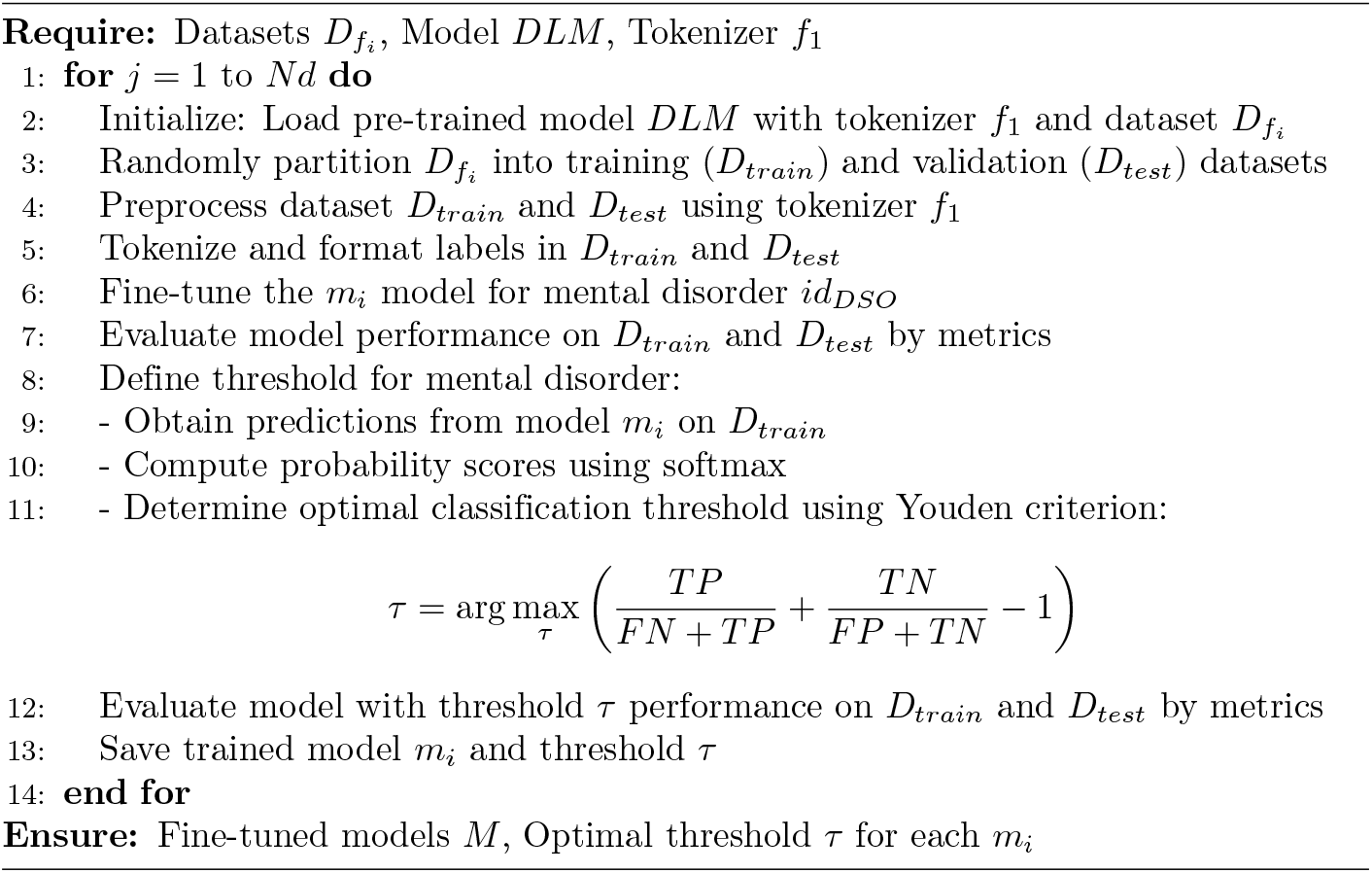

An example of constructing the training dataset *Df*_1_, targeting the mental disorder group *d*_1_, is shown in Figure 2.

**Fig 2.**
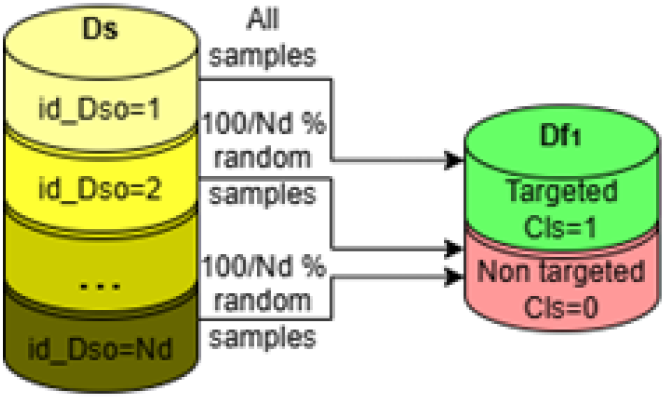
Example of forming dataset *Df*_1_ for disorder *d*_1_ (*id*_*DsO*_ = 1).

Figure 3 presents a data processing diagram for determining the presence or absence of mental disorders.

**Fig 3.**
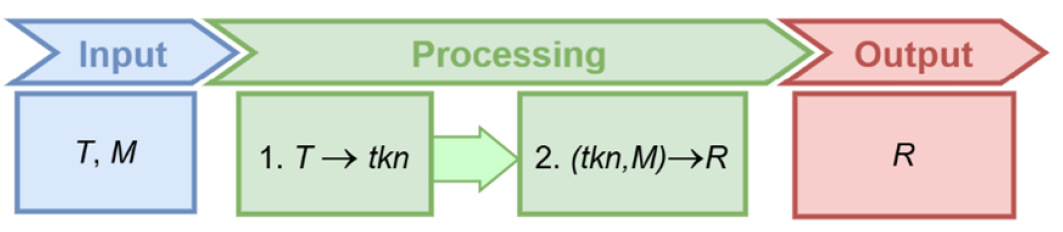
Data processing diagram for determining the presence or absence of a specific mental disorder.

At step 1 (see Fig. 3), the tokenized text *tkn* is obtained from the input text *T* using the mapping *f*_1_. At step 2 (see Fig. 3), Algorithm 3 is used to apply the model *M* to the tokenized text *tkn*, resulting in a decision *R* regarding the presence or absence of mental disorders (mapping *f*_4_).

#### Algorithm 3

Obtaining solution *R* (mapping *f*_4_)

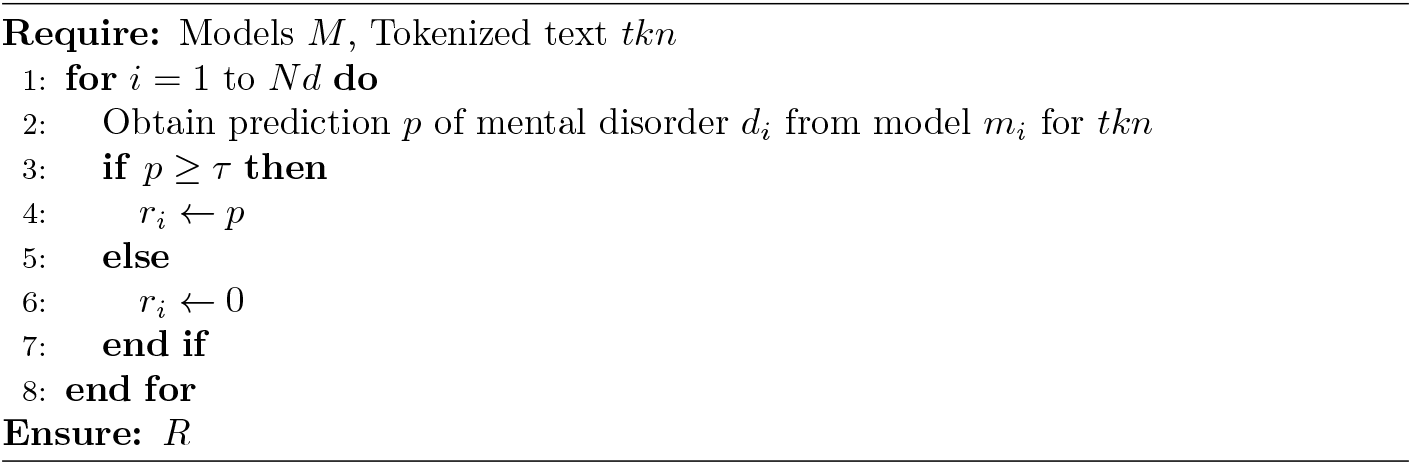

### Datasets

To implement and test the proposed approach, two publicly available datasets from Kaggle were utilized: “Text Classification” [22], [23], and “Depression: Reddit Dataset (Cleaned)” [24]. The study focused on the following five mental disorders: Anger/Intermittent Explosive Disorder (*d*_1_), Anxiety Disorder (*d*_2_), Depression (*d*_3_), Narcissistic Disorder (*d*_4_), and Panic Disorder (*d*_5_). Samples representing these specific disorders were extracted from the “Text Classification” dataset, while samples exhibiting no disorder manifestations (*d*_0_) were sourced from the “Depression: Reddit Dataset (Cleaned)”.

The “Text Classification” dataset consists of 740 tweets referencing potential mental disorders. The data is distributed as follows: 208 samples for *d*_3_ (Depression), 158 for *d*_4_ (Narcissistic Disorder), 154 for *d*_1_ (Anger), 153 for *d*_2_ (Anxiety), and 112 for *d*_5_ (Panic Disorder).

The “Depression: Reddit Dataset (Cleaned)” dataset is a refined collection of 7,650 Reddit posts related to depression. Of these, 3,900 samples show no signs of depression or other mental health issues. This dataset is released under the CC0: Public Domain license, which permits unrestricted public use.

Both datasets contain secondary data which are freely and publicly available, requiring no prior approval. They do not include any personally identifiable or private information. Ethical approval for this research was obtained from the review board of the Faculty of Information Technologies at Khmelnytskyi National University. The research exclusively focused on diagnostic efficacy based on text, with no inclusion of clinical metadata (e.g., age, gender, comorbidities) to ensure confidentiality.

### Experimental Setup and Evaluation

This section details the experimental design, model architecture, training protocol, and evaluation metrics used to validate the proposed approach.

To demonstrate the effectiveness of our approach, we conducted a series of experiments with two primary objectives:

1. To evaluate the capacity of the proposed models to identify textual patterns related to mental disorders and accurately classify those mental disorders based on those patterns.
2. To compare the performance of the proposed approach against other established methods in the field.

The core of our approach is a set of models, *M*, based on the DistilBERT architecture. We chose DistilBERT because standard BERT models can be prone to overfitting on specific words or markers, which may lead to biased conclusions, especially in emotionally charged texts. DistilBERT, with its reduced depth and training via distillation, offers more stable generalization, which is crucial for analyzing texts from diverse individuals.

The datasets utilized in this study present certain limitations, including moderate sample sizes, class imbalance, and potential stylistic and contextual biases inherent to social media data. To mitigate these constraints, we employed a fine-tuning strategy in conjunction with cross-validation. We adopted a 4-fold cross-validation scheme, as research [42] has demonstrated this to be a well-established and robust practice for similar classification tasks. The training and evaluation protocol for each model *m ∈ M* on its corresponding dataset *D*_*f*_ was as follows:

1. *Data Splitting* : The dataset *Df* was randomly split into training and test sets in the proportion of 80% for training and 20% for test, respectively.
2. *Model Fine-Tuning* : The pre-trained DistilBERT model m was fine-tuned on the training set using the following hyperparameters: a batch size of 4, 4 training epochs, and a learning rate of 2e-5. These hyperparameters were selected based on established best practices for transformer-based models on similarly sized datasets. During this process, the fine-tuning loss functions (e.g., focal loss, dice loss, log loss) were monitored.
3. *Evaluation*: The performance of the fine-tuned model was evaluated on both the training and test sets. For the test set, we generated a confusion matrix, calculated performance metrics, and plotted the ROC curve.
4. *Repetition for Robustness*: To ensure the stability and reliability of the results, Steps 1-3 were repeated four times using different random splits of the data.
5. *Result Aggregation*: The final performance was reported as the average of the metrics obtained across all four runs. We also calculated the maximum deviation from the mean to estimate the variance of the results for the *F*_1_-Score metric.

Since the model relies on multi-label classification and accurate mental health diagnosis is critical for human well-being, a comprehensive set of metrics was used to evaluate its performance. These metrics included:

- Classification Metrics: Accuracy, Precision, Recall, and *F*_1_-Score.
- Diagnostic Tools: Confusion Matrices and Receiver Operating Characteristic (ROC) curves.
- Loss Functions: Focal Loss, Dice Loss, and Log Loss were used during training to handle class imbalance and guide the optimization process.

Additionally, the Youden’s *J* Index was employed to determine the optimal threshold for classification, minimizing the risk of both false positives and false negatives, which is crucial in a clinical context.

The experiments were implemented in an IPython Notebook environment. For training the five individual disorder models, CPU execution environments were used, while a TPU v2-8 accelerator was utilized for the multi-class classifiers. The complete source code has been made publicly available on GitHub to ensure reproducibility: https://github.com/oovcharuk/MentalHealthTextClassifier. Per our methodology, Algorithm 1 was used to preprocess and structure the corresponding dataset *D*_*f*_ for each model in *M*.

## Results

After applying Algorithm 1, the following data distribution was obtained in the datasets: *Df*_1_ contains 154 entries for “Anger/Intermittent Explosive Disorder” and 150 entries for the category “non-target”; *Df*_2_ includes 153 entries for “Anxiety Disorder” and 150 entries for “non-target”; *Df*_3_ contains 208 entries for “Depression” and 205 entries for “non-target”; *Df*_4_ includes 158 entries for “Narcissistic Disorder” and 155 entries for “non-target”; *Df*_5_ consists of 112 entries for “Panic Disorder” and 110 entries for “non-target”.

After completing step 1 (Experiment for evaluating *M* models), the following distributions of training and test data were obtained: for *d*_1_, the training set contained 243 samples (80%) and the test set 61 (20%); for *d*_2_, training: 242 (80%), test: 61 (20%); for *d*_3_, training: 330 (80%), test: 83 (20%); for *d*_4_, training: 250 (80%), test: 63 (20%); for *d*_5_, training: 177 (80%), test: 45 (20%).

At step 2, a set of models *M* was experimentally trained for each of the 5 types of psychological disorders. In step 3, the loss function values were obtained for each model. In step 4, the aforementioned procedures were replicated using four random 80/20 splits. The rationale for selecting four splits is detailed in the “Experimental Setup and Evaluation” section. In step 5, the average values of all metrics were obtained, and the maximum deviation from the average value was calculated for the *F*_1_-score metric.

Table 2 presents the mean values for Focal Loss, Dice Loss, and Log Loss across both the training and test datasets. The integration of these loss functions allows the model to simultaneously address class imbalance, optimize the *F*_1_-score, and ensure training stability. Consequently, the values reported in Table 2 provide a representative assessment of the model’s behavior under various scenarios.

**Table 2.**
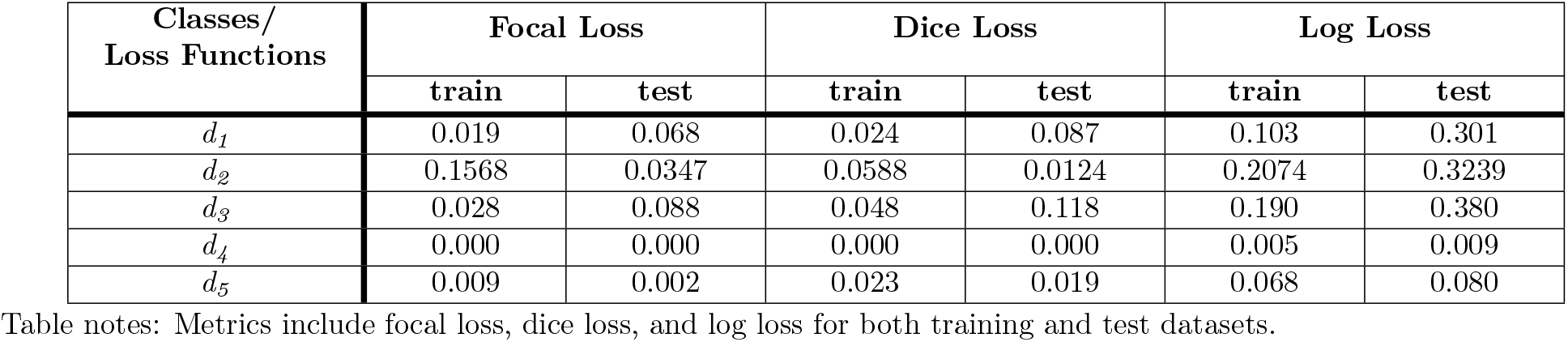
The value of loss functions for binary models with *M*.

| Classes/<br>Loss Functions | Focal Loss |  | Dice Loss |  | Log Loss |  |
| --- | --- | --- | --- | --- | --- | --- |
|  | train | test | train | test | train | test |
| $d_1$ | 0.019 | 0.068 | 0.024 | 0.087 | 0.103 | 0.301 |
| $d_2$ | 0.1568 | 0.0347 | 0.0588 | 0.0124 | 0.2074 | 0.3239 |
| $d_3$ | 0.028 | 0.088 | 0.048 | 0.118 | 0.190 | 0.380 |
| $d_4$ | 0.000 | 0.000 | 0.000 | 0.000 | 0.005 | 0.009 |
| $d_5$ | 0.009 | 0.002 | 0.023 | 0.019 | 0.068 | 0.080 |

Table notes: Metrics include focal loss, dice loss, and log loss for both training and test datasets.

Table 3 shows the average values of the statistical metrics Accuracy, Precision, Recall, and *F*_1_-score for the training and test data. The Delta *F*_1_ column contains the maximum deviation from the average value for the *F*_1_-score metric.

**Table 3.**
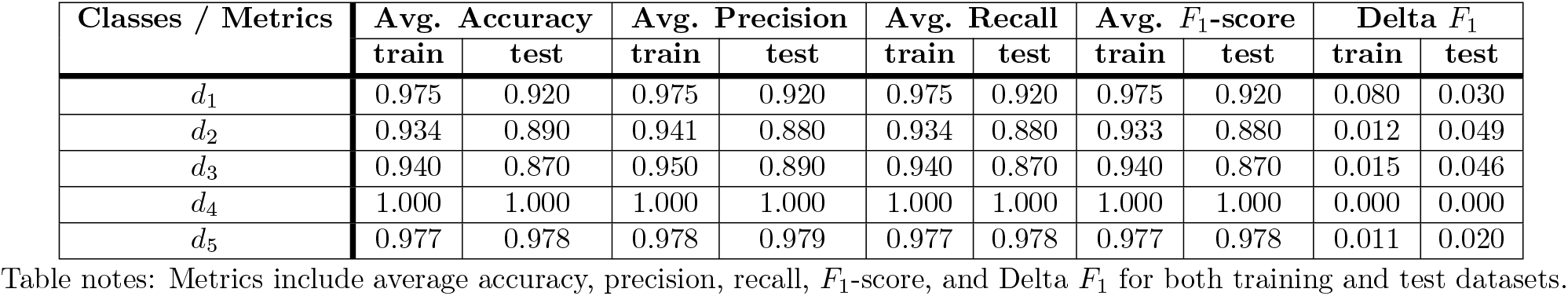
Metrics of binary models with *M* on training and test data.

| Classes / Metrics | Avg. Accuracy | | Avg. Precision | | Avg. Recall | | Avg. $F_1$-score | | Delta $F_1$ | |
| --- | --- | --- | --- | --- | --- | --- | --- | --- | --- | --- |
|  | train | test | train | test | train | test | train | test | train | test |
| $d_1$ | 0.975 | 0.920 | 0.975 | 0.920 | 0.975 | 0.920 | 0.975 | 0.920 | 0.080 | 0.030 |
| $d_2$ | 0.934 | 0.890 | 0.941 | 0.880 | 0.934 | 0.880 | 0.933 | 0.880 | 0.012 | 0.049 |
| $d_3$ | 0.940 | 0.870 | 0.950 | 0.890 | 0.940 | 0.870 | 0.940 | 0.870 | 0.015 | 0.046 |
| $d_4$ | 1.000 | 1.000 | 1.000 | 1.000 | 1.000 | 1.000 | 1.000 | 1.000 | 0.000 | 0.000 |
| $d_5$ | 0.977 | 0.978 | 0.978 | 0.979 | 0.977 | 0.978 | 0.977 | 0.978 | 0.011 | 0.020 |

Table notes: Metrics include average accuracy, precision, recall, $F_1$ -score, and Delta $F_1$ for both training and test datasets.

The ROC curves and confusion matrices for each disorder are presented below. It is important to note that these visualizations depict a single, representative experimental run, whereas the metrics provided in Table 3 are averaged across four data splits.

For “Anxiety Disorder” (*d*_2_), the optimal threshold – determined via Youden’s index as described in the “Materials and methods” section – is 0.7401; the corresponding confusion matrix and ROC curve are shown in Figure 4.

**Fig 4.**
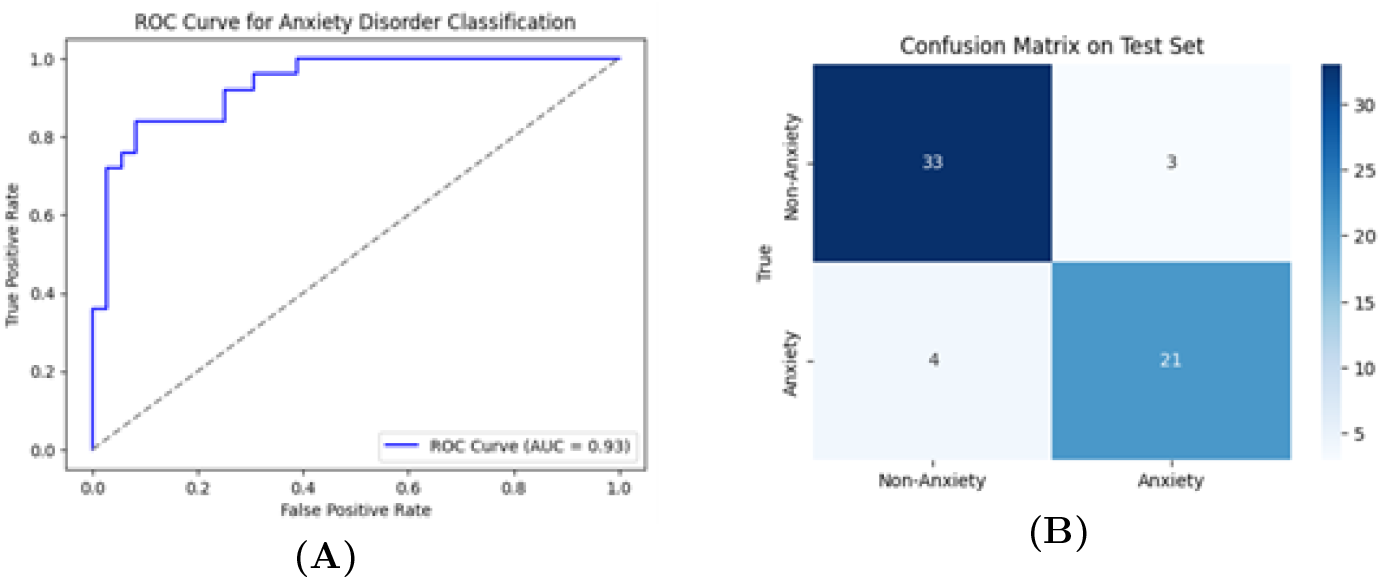
Anxiety disorder class diagrams: A: ROC curve for “Anxiety disorder” classification. B: Confusion matrix on test set.

**Fig 5.**
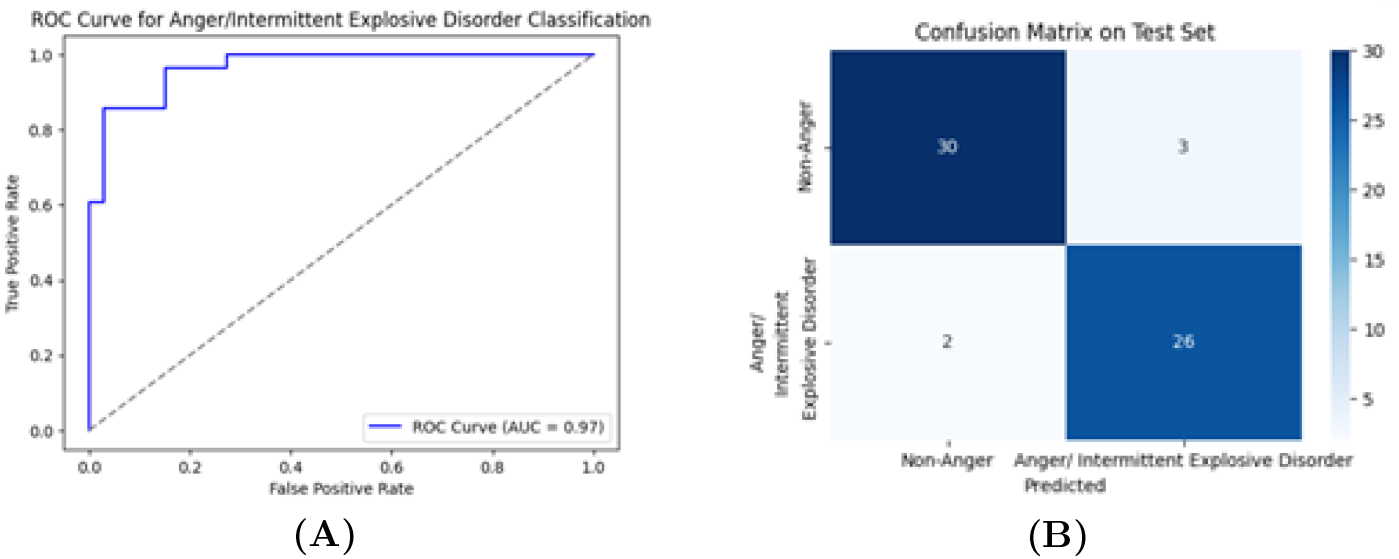
Anger/Intermittent explosive disorder class diagrams: A: ROC curve for “Anger/Intermittent explosive disorder” classification. B: Confusion matrix on test set.

The optimal threshold for “Depression” (*d*_3_) is 0.501. The confusion matrix and ROC curve are shown in Figure 6.

**Fig 6.**
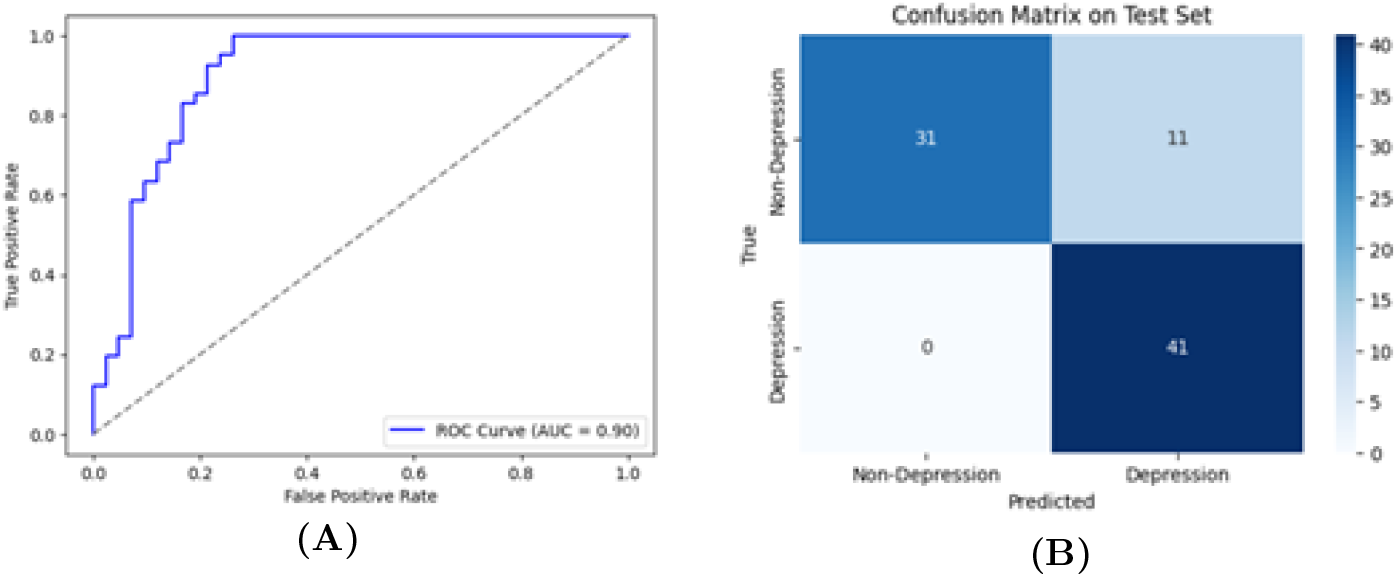
Depression class diagrams: A: ROC curve for “Depression” classification. B: Confusion matrix on test set.

For “Panic Disorder” (*d*_5_), the optimal threshold is 0.6094. The confusion matrix and ROC curve are shown in Figure 7.

**Fig 7.**
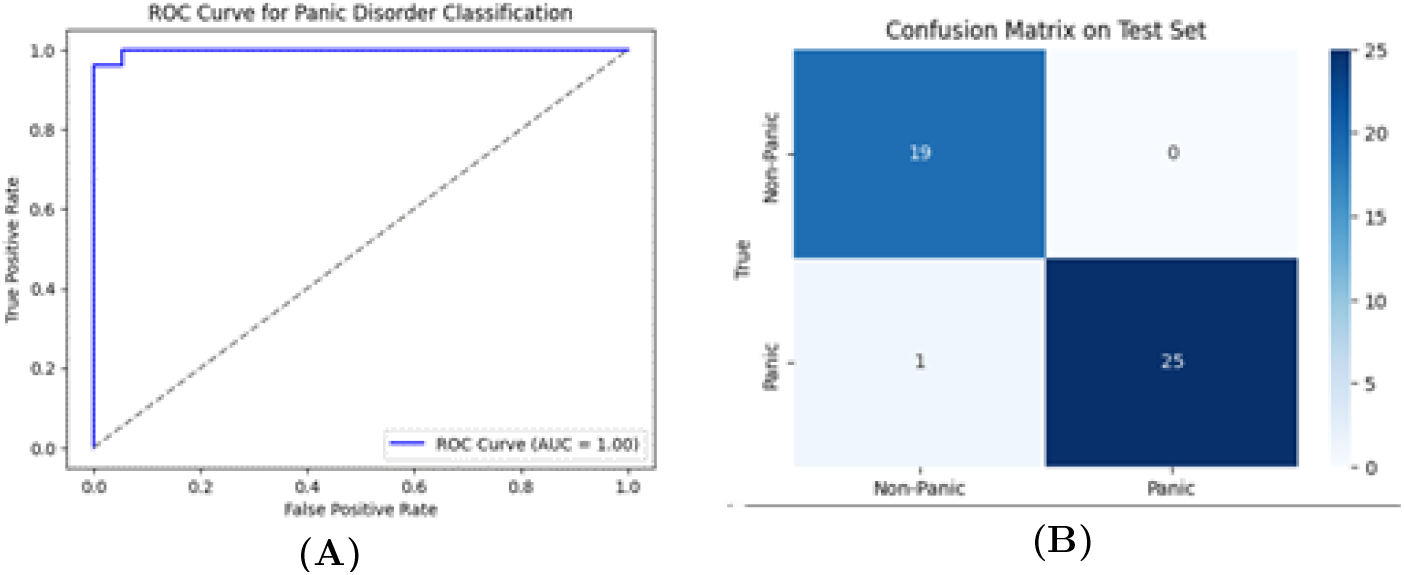
Panic Disorder class diagrams: A: ROC curve for “Panic Disorder” classification. B: Confusion matrix on test set.

For “Narcissistic Disorder” (*d*_4_), the optimal threshold is 0.8978. The confusion matrix and ROC curve are shown in Figure 8.

**Fig 8.**
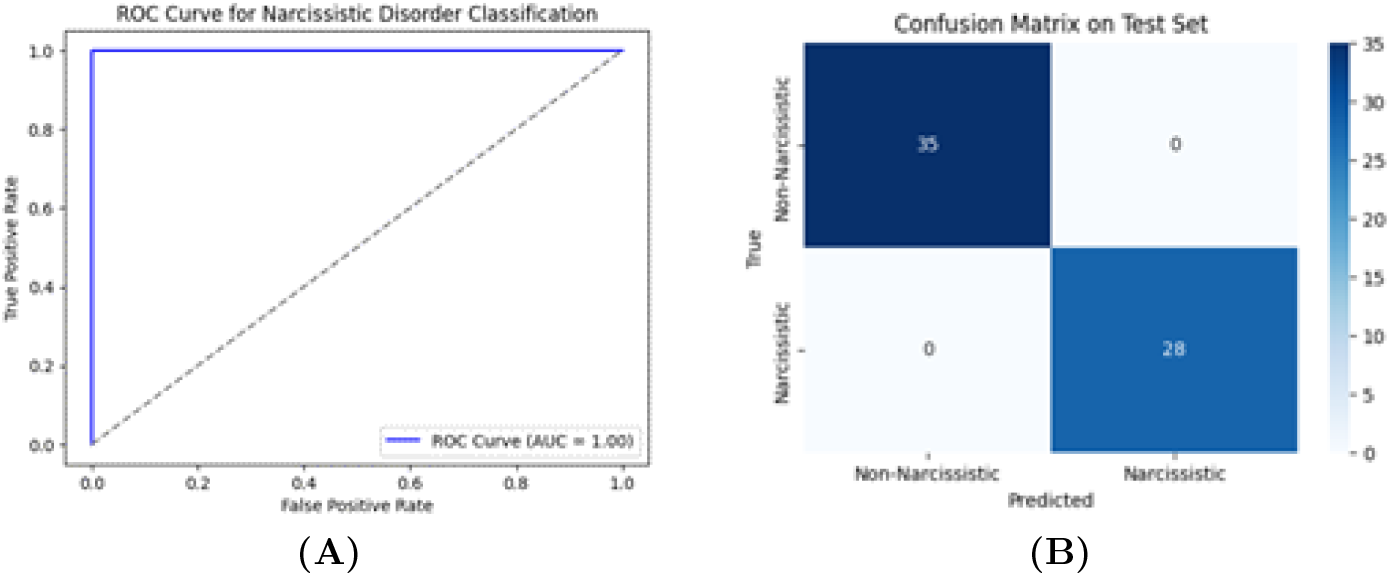
Narcissistic Disorder class diagrams: A: ROC curve for “Narcissistic Disorder” classification. B: Confusion matrix on test set.

Table 4 contextualizes the performance of the proposed ensemble framework against benchmarks reported in recent state-of-the-art studies. While a direct side-by-side comparison is precluded by the unavailability of specific external datasets used in [16] and [19], this overview highlights the efficacy of the proposed binary relevance strategy relative to traditional multi-class architectures employed in similar mental health monitoring tasks.

**Table 4.**
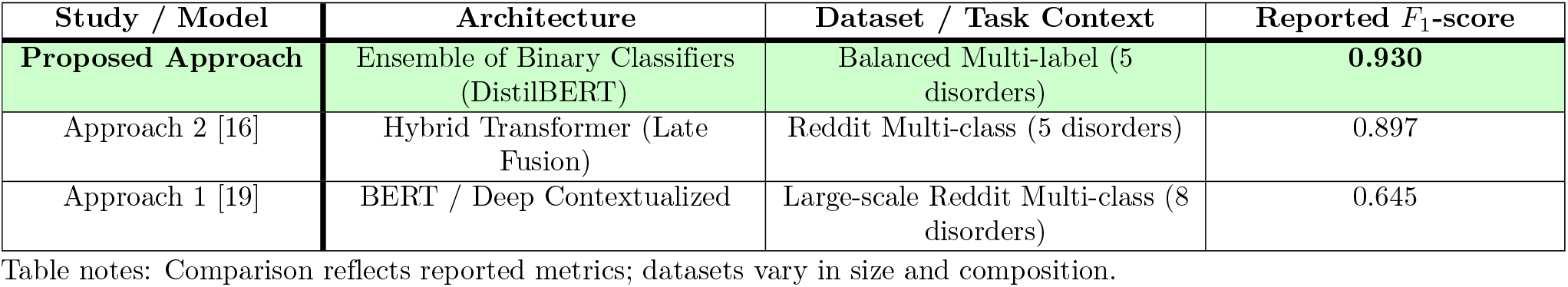
Performance overview of the proposed method versus reported baselines.

## Discussion

In this study, we presented a transformer-based ensemble framework for the multi-label detection of mental health disorders. The proposed approach achieved a high overall *F*_1_-score of 0.930, with AUC values consistently exceeding 0.89 across all modeled conditions. These results suggest that decomposing the multi-class problem into independent binary relevance tasks, combined with threshold optimization via Youden’s *J* statistic, effectively captures the linguistic nuances of co-occurring mental health conditions.

### Impact of Loss Functions

The choice of loss function proved critical for model performance. Our experiments indicate that Focal Loss is particularly effective for hard-to-classify or minority classes, such as “Panic Disorder”, as it penalizes difficult examples more heavily than standard Cross-Entropy. Conversely, Dice Loss demonstrated strong generalization capabilities for classes with ambiguous boundaries, such as “Anxiety Disorder”. The combination of these functions allowed the ensemble to mitigate the effects of class imbalance inherent in social media datasets, yielding stable convergence where “standard” Log Loss underperformed.

### Linguistic Interpretation and Error Analysis

A qualitative analysis of misclassifications reveals that while the models exhibit high sensitivity, they are prone to specific linguistic pitfalls that vary by disorder:

- **Contextual Polarity Shifting (Anxiety):** False negative errors in the “Anxiety Disorder” model often stemmed from polarity shifts induced by negations and modals. Statements such as *“I am paralyzed by fear and cannot make decisions”* or *“I am worried that I cannot cope with my anxiety”* were misclassified because the negation markers (“cannot”, “not”) locally inverted the sentiment intensity in the embedding space. Conversely, false positives were triggered by “panic spectrum” vocabulary in non-target contexts (e.g., *“I isolate myself so as not to cause panic”*), where the combination of worry and avoidance amplified the predictive signal despite the lack of clinical intent.
- **Metaphorical Ambiguity (Anger/IED):** The “Anger” model showed a slight tendency toward over-prediction driven by metaphorical descriptions. Non-target instances like *“my body feels like a pressure cooker, ready to explode”* contain aggregated keywords (“pressure”, “explode”) that semantically align with explosive anger, generating false positives. On the other hand, euphemistic phrasing (e.g., *“Trying to find ways to manage emotions without them managing me”*) attenuated the signal, leading to false negatives.
- **Colloquial vs. Clinical Usage (Depression):** While the “Depression” model achieved near-perfect sensitivity (0 False Negatives in the representative run), specificity was impacted by the colloquial use of depressive terminology. False positives were concentrated in statements where keywords like “sad” or “depressed” referred to external events or third parties (e.g., *“fadyanwar is sad because it was the last GSM company”* or references to celebrities like *“David Archuleta”*). In these cases, the model acted on superficial triggers without capturing the lack of personal symptomatic distress.
- **Dataset Constraints (Narcissistic Disorder):** The perfect classification of “Narcissistic Disorder” (0 FP, 0 FN) at a high threshold (*τ* =0.8978) likely indicates distinct linguistic markers in this subset, but may also suggest limited variability in the training data, posing a risk of overfitting to specific keywords.

### Comparison with Benchmarks

Our binary ensemble approach compares favorably with existing benchmarks. Our mean *F*_1_-score (0.930) surpasses the results reported in related studies using standard multi-class BERT architectures (*F*_1_*≈*0.65–0.73) [19] and hybrid deep learning models (*F*_1_*≈*0.89) [16]. Unlike traditional multi-class classifiers that force a single label per text, our multi-label framework acknowledges the comorbidity of mental disorders, providing a more realistic modeling of user states. The comparison suggests that shifting from a multi-class paradigm to a set of independent binary tasks significantly reduces inter-class confusion, leading to higher *F*_1_-scores on targeted datasets.

### Limitations and Future Work

Several limitations must be acknowledged to contextualize these findings. First, the relatively small training corpus raises concerns about potential overfitting, particularly for classes with high performance like Narcissistic Disorder. Second, the ground truth labels are derived from user-generated content and self-reports rather than clinical diagnoses. Consequently, the model detects discourse patterns associated with disorders, rather than the disorders themselves. Finally, the available data lacks demographic metadata (such as age, gender, or location), which prevents the assessment of fairness or potential bias across different population groups.

The deployment of AI in mental health requires strict ethical oversight. The identified potential for false positives, driven by colloquialisms and metaphors, underscores that such systems should function solely as decision support tools for professionals, not as autonomous diagnostic agents. Future research will focus on expanding the dataset to include more diverse sources, integrating explainer algorithms (e.g., SHAP) to improve interpretability, and refining the handling of negations and irony to reduce linguistic misclassifications.

## Conclusion

This study presented a novel multi-label classification framework for mental health monitoring, integrating an ensemble of fine-tuned DistilBERT models with a calibrated binary relevance strategy. By decomposing the diagnostic task into independent detection problems, the proposed approach had a mean *F*_1_-score of 0.930. These results indicate that replacing traditional multi-class architectures with specialized binary classifiers significantly reduces confusion between clinically overlapping conditions, such as depression and anxiety, thereby offering a more effective method for modeling comorbidity in social media text.

Beyond quantitative metrics, this research provided a critical qualitative analysis of the linguistic boundaries of transformer-based models. We have identified that while the proposed approach has high sensitivity, specificity has been challenged by contextual polarity shifting (e.g., negations), metaphorical ambiguity, and the colloquial usage of clinical terms. These findings highlight that it can still be challenging for high-performing neural networks to capture all the semantic nuances required for precise diagnosis, which highlights the existing gap between statistical correlation and language specifics.

In summary, while the proposed framework demonstrates significant potential as a screening and monitoring tool, it must be deployed and used with strict human oversight to mitigate the risks of false positives driven by linguistic artifacts. Potential future research directions will focus on enhancing model interpretability through explainer algorithms, expanding datasets to include diverse demographic and cross-cultural samples, and developing architectures capable of better resolving syntactic dependencies to minimize errors caused by negation, irony, and colloquial language.

## Data Availability

All data presented in this paper are contained in the manuscript and in references provided in the manuscript.

https://www.kaggle.com/datasets/comsys/text-classification

https://www.kaggle.com/datasets/infamouscoder/depression-reddit-cleaned

https://github.com/oovcharuk/MentalHealthTextClassifier

## Author contributions

**Conceptualization:** Olexander Barmak, Iurii Krak, Pavel Skums, Sergiy Yakovlev.

**Data curation:** Oleksandr Ovcharuk, Maryna Molchanova.

**Formal analysis:** Oleksandr Ovcharuk, Maryna Molchanova.

**Project administration:** Olexander Barmak, Sergiy Yakovlev.

**Investigation:** Oleksandr Ovcharuk, Olexander Mazurets.

**Methodology:** Oleksandr Ovcharuk, Olexander Mazurets.

**Software:** Oleksandr Ovcharuk, Maryna Molchanova.

**Supervision:** Olexander Mazurets.

**Validation:** Olexander Mazurets, Alexander Kirpich, Pavel Skums.

**Writing – original draft:** Oleksandr Ovcharuk, Maryna Molchanova.

**Writing – review & editing:** Olexander Mazurets, Alexander Kirpich, Pavel Skums, Olena Sobko, Sergiy Yakovlev.

## Notes

### Competing Interest Statement

The authors have declared no competing interest.

### Funding Statement

This study did not receive any funding

### Author Declarations

All data are fully available without restriction: The datasets underlying the results of this study are available on Kaggle: the Text Classification dataset (https://www.kaggle.com/datasets/comsys/text-classification) and the Depression: Reddit Dataset (Cleaned) (https://www.kaggle.com/datasets/infamouscoder/depression-reddit-cleaned). The source code and pre-processing scripts are available on GitHub at https://github.com/oovcharuk/MentalHealthTextClassifier.

